# Gender score development in a retrospective approach in the Berlin Aging Study II

**DOI:** 10.1101/2019.12.16.19015032

**Authors:** A. Tauseef Nauman, Hassan Behlouli, Nicholas Alexander, Friederike Kendel, Johanna Drewelies, Konstantinos Mantantzis, Nora Berger, Gert G. Wagner, Denis Gerstorf, Ilja Demuth, Louise Pilote, Vera Regitz-Zagrosek

## Abstract

In addition to biological sex, gender, the sociocultural dimension of being a woman or a man, plays a central role in health. However, there are so far no approaches to quantify gender in a retrospective manner in existing study datasets. We therefore aimed to develop a methodology that can be retrospectively applied to assess gender in existing cohorts. We used baseline data from the Berlin Aging Study II (BASE-II), obtained in 2009-2014 from 1869 participants aged 60 years and older. We identified 13 gender related variables and used them to construct a gender score (GS) by primary component and logistic regression analysis. Of these, 9 variables contributed to a gender score: chronic stress, marital status, risk taking behavior, agreeableness, neuroticism, extraversion, loneliness, conscientiousness, and education. GS differed significantly between females and males as defined by sex. Next, we calculated linear regressions to investigate associations between sex, GS, and selected biological and well-being variables. Sex, but not GS was significantly associated with LDL-C and TC. GS, but not sex, was significantly associated with cortisol levels, CES-depression, negative affect, life satisfaction. Thus, we were able to develop a GS in a retrospective manner from available study variables that characterized women and men in addition to biological sex. This approach will allow us to introduce the notion of gender retrospectively into a large number of studies.

## Introduction

Biological sex and gender, the sociocultural dimension of being a woman or a man, play a central role in health.^1-4^ Unlike sex, which is biologically based, gender incorporates the sociocultural dimension of being a woman or a man in a given society.^1, 2^ It determines interaction of patients with doctors and the health care system, help-seeking behavior and access to care. Since biological sex and gender overlap, but are not identical, gender is likely to influence health differently from biological sex.^1, 2^ Whereas sex is relatively easy to quantify, few possibilities exist to quantify gender in a reproducible manner.

Recently, the Canadian members of our group and their partners constructed a gender score (GS) in a prospective manner. This GS included a variety of carefully selected psychosocial and sociocultural variables, in order to characterize and distinguish women and men from one another over and above biological sex in cohort studies.^1, 2^ In a prospective study in patients with ACS, the GS was more strongly associated with several important cardiovascular (CV) risk factors and one-year mortality than biological sex. ^1, 2^

It was concluded that the GS can be used to evaluate the effect of gender independent from the effects of sex on disease presentation, processes of care, and relevant outcomes. Including such a gender dimension into clinical studies may thus add new explanatory value to understand clinical outcomes. Therefore, it would be desirable to calculate a GS in as many study databases as possible.

Since the majority of existing studies did not include a GS in a prospective manner, we aimed in the present study to develop a methodology that could be retrospectively applied so as to assess gender in existing study cohorts. This should be possible since most study databases include a large number of psychosocial and/or socioeconomic variables that are potentially gender-related and could be used to construct a GS. We therefore decided to use, in a paradigmatic approach, the Berlin Aging Study II (BASE-II) to develop a strategy to construct a GS in a retrospective manner. We used the data, obtained in BASE-II from 2009-2014, and selected potentially gender-related variables from the initial investigation that covered the different dimensions of gender and that were as close as possible to those included in the prospectively developed score from GENESIS-PRAXY. We then tested the distribution of this score between women and men and analyzed its independent associations with clinical disease indicators and biological risk factors.

## Methods

We used baseline data from the Berlin Aging Study II (BASE-II), obtained in 2009-2014 from 1869 participants aged 60 years and older.^5^ Because of the larger BASE-II sample of n=2200 participants was tested in different years from different research units, not all data were available in all cases and we focused on those data available in more than 65 % of cases.

First we selected potentially gender related variables from the study database that reflect different aspects of the gender construct (e.g. gender roles, gender typical behavior) in which women and men traditionally differ.^6, 7^ These variables were selected to replicate as close as possible the items used in the GENESIS-PRAXY study, related to social roles and social support related variables (employment status, family status, level of Education) as well as gender related personality traits: chronic stress (assessed using the Trier Chronic Stress Inventory, TICS); perceived stress (assessed using the Perceived Stress Scale, PSS)^8^ loneliness (assessed using the UCLA Loneliness Scale)^9^; Big five personality traits (using the BIG Five Inventory with its dimensions “openness to experience”, “conscientiousness”, “extraversion”, “agreeableness” and “neuroticism”)^10^; risk-taking behavior^11^. All were measured using validated self-report questionnaires.

When possible, we calculated scores according to questionnaire standard. For the BFI, TICS, and UCLA-Loneliness, mean scores across items were used (for details, see Gerstorf et al., 2015; Kühn, Düzel, Drewelies, Gerstorf, Lindenberger, & Gallinat, 2018).^12, 13^

All participants gave written informed consent. The Ethics Commission of the Berlin Chamber of Physicians (Ärztekammer Berlin) approved the BASE study prior to the first assessments in 1990 (the approval was not numbered), and the Ethics Committee of the Charité – Universitätsmedizin Berlin approved the BASE-II study (approval number EA2/029/09)^14^.

### Overall strategy and statistical approach

Our approach was divided in 3 steps (figure 1): GS development, GS calculation in individual cases and GS application.

**Figure 1.**
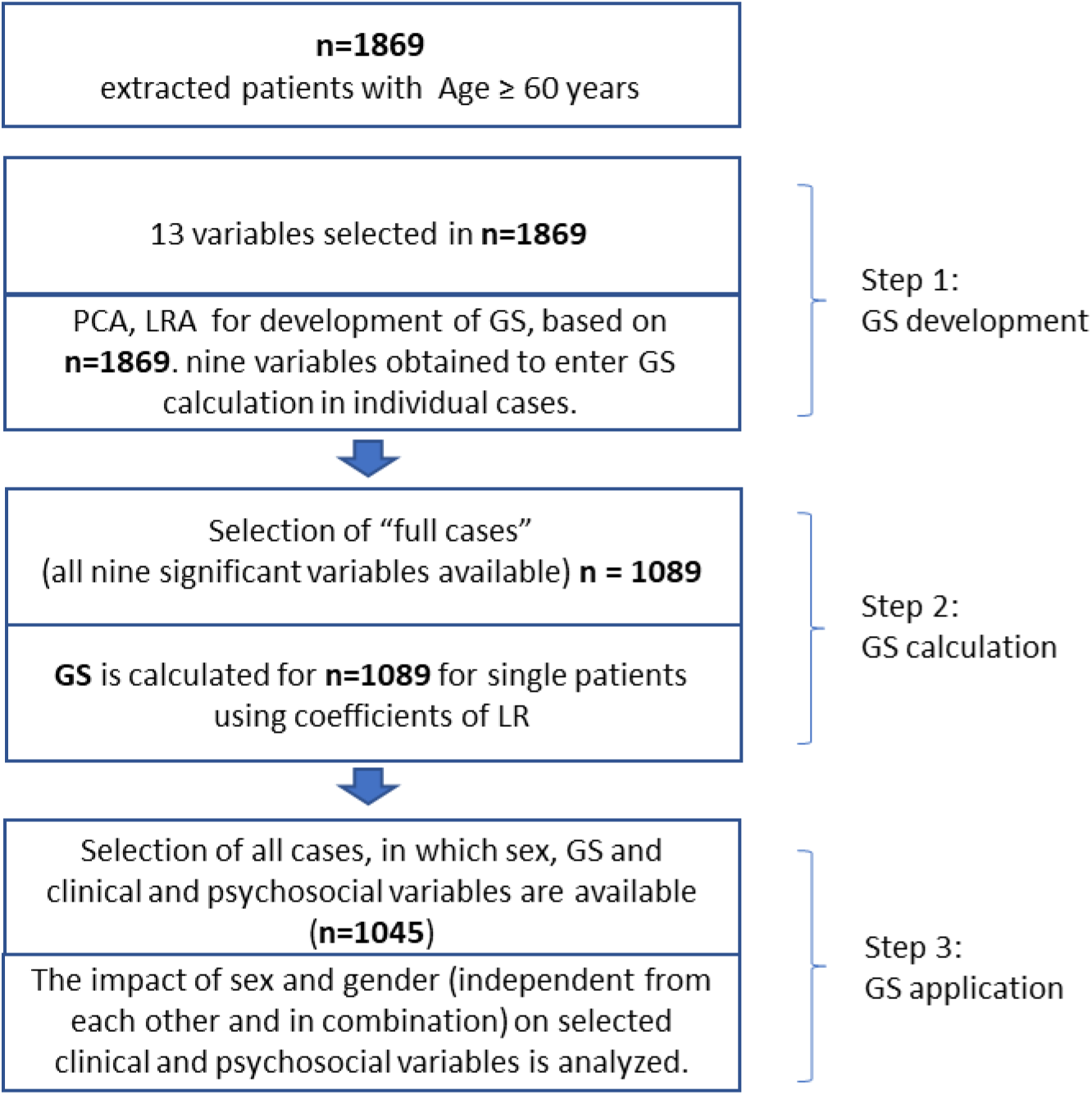
Stratification of data analysis. Stratification of data analysis was divided into three major steps. Where in step 1 and 2 Gender Score (GS) was developed and Step3 represents the implication of the GS.

#### GS Development

For step 1 all 1869 cases were used. First, a bivariate correlation analysis was performed and for each significantly correlated pair of variables with a correlation coefficient equal or greater than 0.80, one of the two variables was randomly removed. Next, PCA analysis was conducted with a varimax rotation method to validate the component matrix results. In interpreting the factor analysis, an item was said to load on a given component if the factor loading was 0.40 or greater for that component and was less for other components.

Based on the remaining variables, Logistic regression (LR) models for the association with sex were calculated one by one and all non-significant variables were removed in a descending order based on the size of their non-significant p-value.

#### GS calculation in individual cases

For step 2, the coefficient estimates of these variables associated with sex were used for GS calculation case by case. For this step, the presence of all variables was required in each study subject. This reduced the number of cases available for calculation of individual GS to 1089 (Fig 1).

#### GS Application

In a 3^rd^ step, the association of GS with clinical and psychosocial variables was tested using parametric and non-parametric tests as well as linear regression analyses. This step required the presence of GS and the clinical/ psychosocial investigation in all cases. This reduced the number of cases to 1045.

#### Statistical tests

SPSS version 25 was used to perform statistical analysis. For the comparison of means Chisq. Test or T-test and Mann-Whitney U test was used where appropriate. Bivariate correlations were calculated with 0.80 as the cut off values. Logistic models were used to model the probability of associated variables. To look for an index of sensitivity and specificity the Curve statistic was performed for GS (0-100) as the test variable and sex (0 = men, 1 = women). Significance level was determined for p < 0.05 for the group differences and LR models.

## Results

### Development of the gender score (GS)

To calculate the GS in the BASE II cohort, we considered a total of 13 variables (table 1). In the first step, we estimated a correlation matrix to test for the existence of highly correlated variables. Since none of the correlations exceeded *r* = 0.58 (Table S1), we retained all 13 variables for the next step of PCA, assessment of factor loading. The 13 variables loaded onto five components (Table 2, <u>PCA, factor loadings).</u>

**Table 1.**
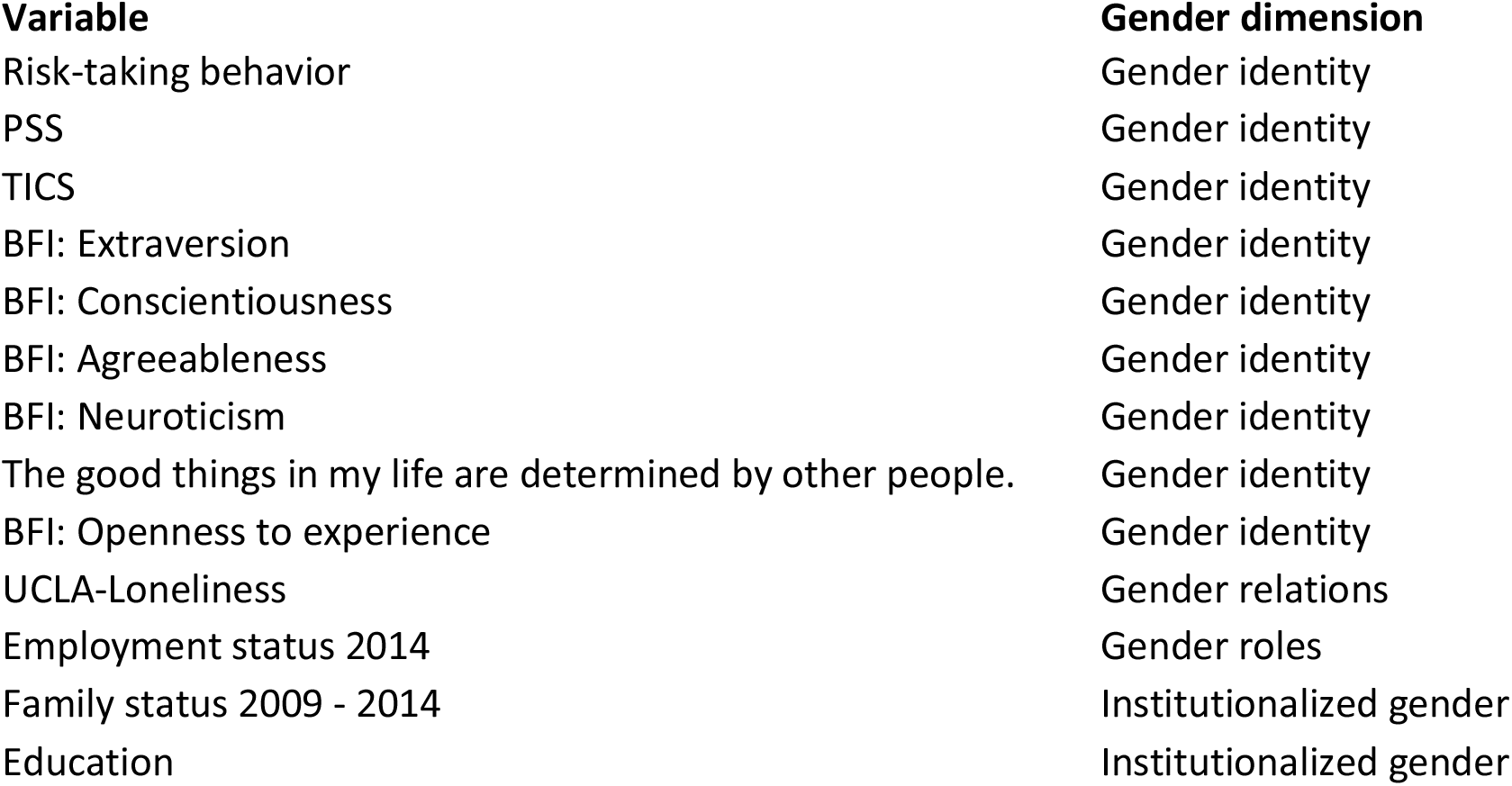
Selected gender related variables in the BASE-II cohort. Each variable was taken according to the definition provided in the Women health research network^7^.

| **Variable** | **Gender dimension** |
| --- | --- |
| Risk-taking behavior | Gender identity |
| PSS | Gender identity |
| TICS | Gender identity |
| BFI: Extraversion | Gender identity |
| BFI: Conscientiousness | Gender identity |
| BFI: Agreeableness | Gender identity |
| BFI: Neuroticism | Gender identity |
| The good things in my life are determined by other people. | Gender identity |
| BFI: Openness to experience | Gender identity |
| UCLA-Loneliness | Gender relations |
| Employment status 2014 | Gender roles |
| Family status 2009 - 2014 | Institutionalized gender |
| Education | Institutionalized gender |

**Table 2.**
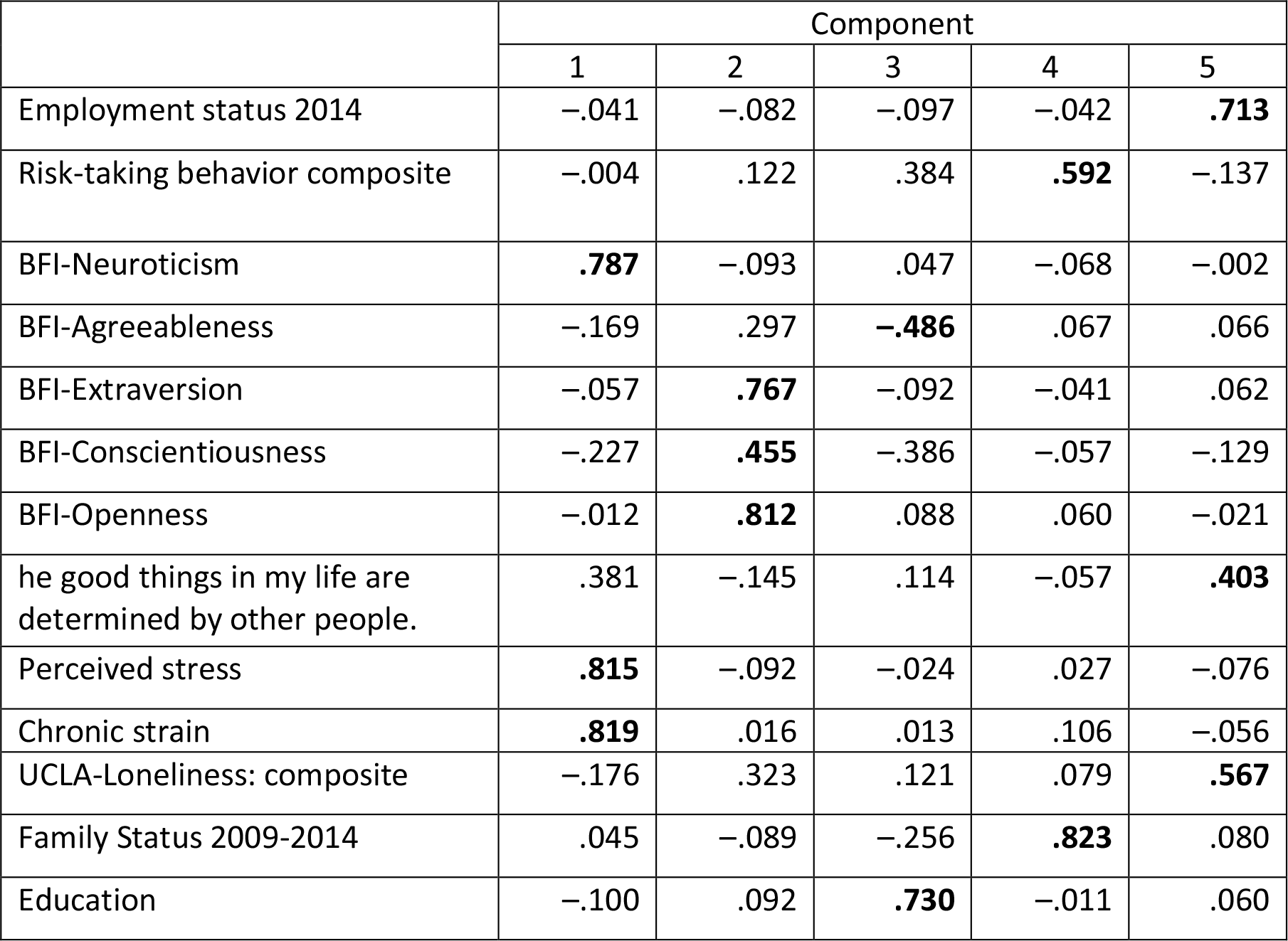
Gender related variables and corresponding factor loading in PCA. In this factor analysis, an item was said to load on a given component if the factor loading was 0.40 or greater for that component and was less than 0.4 for other components. Based on this analysis, all items were included in LR.

In the next step, the 13 variables were used to perform a LR analysis with sex as a dependent variable. Non-significant variables were removed one by one in a descending order of their p-value (0.05). In total, 5 models were explored, the final model is presented in Table 3. The nine statistically significant variables were retained. They were: chronic stress (TICS mean), marital status, risk taking behavior, BFI agreeableness, BFI neuroticism, BFI extraversion, loneliness (UCLA-Loneliness mean), BFI Conscientiousness, and education.

**Table 3.** Significant variables for the calculation of GS. LR Models using the identified variables from PCA were calculated and non-significant variables were removed one by one in a descending order. Finally, the above model was reached with nine independently significant variables. The Area Under the Curve statistic was .795, p < .001, 95% CI [.769, .821] which indicates fair-to-good sensitivity/specificity (test values ranged between 0.5 and 1).

| Variable name | Coefficient estimates | *p* |
| --- | --- | --- |
| TICS | 0.63 | <0.001 |
| Family Status 2009-2014 | 0.52 | <0.001 |
| Risk-taking behaviour | 0.37 | <0.001 |
| BFI: Agreeableness | 0.31 | <0.001 |
| BFI: Neuroticism | 0.27 | <0.001 |
| BFI: Extraversion | 0.27 | <0.001 |
| UCLA-Loneliness | 0.44 | <0.05 |
| BFI: Conscientiousness | 0.17 | <0.05 |
| Education | -0.04 | <0.05 |

### Calculation of gender score (GS) in individual cases

The coefficients of the final logistic regression analysis were used to calculate the gender score (GS), with a range from 0 to 100. Our whole cohort of participants evenly spread over the score. The GS in the 1^st^, 2^nd^, 3^rd^tertile ranged from 2-37, 38-68, 69-98 %, for women and men (table S 2). However, the frequency distribution of women and men differed. 55 % of men were located in the first tertile with more masculine GS and 51 % of women in the 3^rd^tertile with the more feminine scores. At the same time, the distributions showed considerable overlap with a 36% and 31% of women and men in the middle tertile and 13 % of women and 14 % of men having score values of the opposite gender (Fig 2, table S2).

**Figure 2.**
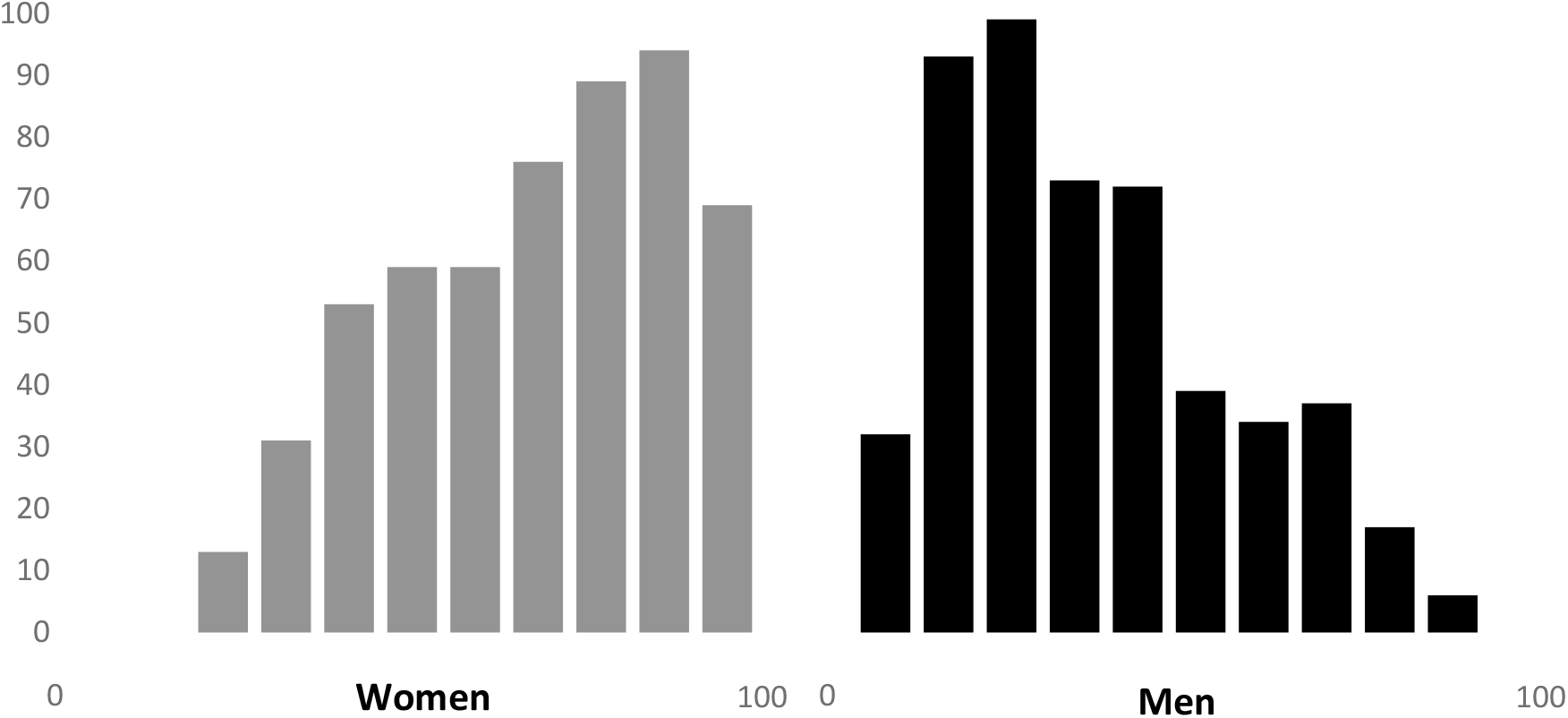
Gender score distribution in women and men. **Distribution of calculated gender score in women and men.** Zero is identical with completely masculine characteristics whereas 100 represents completely feminine characteristics.

To look how well GS separated women and men, a ROC analysis with 1089 participants was performed that had a gender score available. The Area Under the Curve statistic was .795, p < .001, 95% CI [.769, .821] which indicates fair-to-good sensitivity/specificity (test values ranged between 0.5 and 1).

### Association of the GS with clinical and psychosocial variables

Next, we tested the association of the GS with clinical parameters and measures of well-being, including lipid levels, total cholesterol (TC), HbA1c, CES-Depression, cortisol levels, grip strength, negative affect, and life satisfaction. Table 4 and Table 5 show the distribution of these variables according to biological sex and GS tertiles. Most of the variables with significant differences between women and men (Table 4a) were also significantly different across GS tertiles (Table 4b). Body mass index and family history of cardiovascular disease were exceptions, with significant differences when analyzed by sex, but not by gender.

**Table 4 a:** Differences on participant characteristics by biological sex.

|  | **Men (N=502)** | **Women (N=543)** | **p-value<sup>1</sup>** |
| --- | --- | --- | --- |
| Age in years, mean (SD) | 68.8 (3.5) | 68.4 (3.4) | 0.068 |
| Education, median (IQR) | 15 (6) | 13 (6.5) | <0.001 |
| LDL-C (mg/dl), mean (SD) | 124.8 (34.3) | 137.4 (34.3) | <0.001 |
| TC (mg/dl), mean (SD) | 203.8 (38.1) | 227.3 (38.1) | <0.001 |
| BMI, mean (SD) | 27.2 (3.6) | 26.5 (4.7) | 0.004 |
| Blood pressure, systolic (mmHg), mean (SD) | 143.3 (18.3) | 146.2 (58.9) | 0.309 |
| Blood pressure, diastolic (mmHg), mean (SD) | 83.2 (10.9) | 86.0 (60.5) | 0.325 |
| Current smoking (N) | 58 | 38 | 0.013 |
| HbA1c (%), mean (SD) | 5.7 (0.6) | 5.6 (0.6) | 0.012 |
| Family history of CVD (N) | 207 | 264 | 0.044 |
| CES-Depression (sum score), median (IQR) | 5 (7) | 6 (8) | <0.001 |
| Cortisol (nmol/L) mean (SD) | 405.5 (109.6) | 376.4 (114.7) | <0.001 |
| Hand grip strength (kg), mean (SD) | 42.6 (6.8) | 26.5 (4.8) | <0.001 |
| Negative affect, mean (SD) | 47.7 (9.3) | 48.7 (9.2) | 0.080 |
| Life satisfaction, median (IQR) | 53.3 (11.7) | 49.4 (15.6) | 0.003 |

<sup>1</sup>Differences between men and women were assessed using the parametric t test or the nonparametric Mann-Whitney U test where appropriate. The $\chi^2$ test was used to determine the difference in current smoking and family history of CVD between men and women. <sup>2</sup> Number of available data. Abbreviations: LDL-C, low density lipoprotein-cholesterol; TC, total cholesterol; BMI, body mass index; HbA1c, glycated hemoglobin A1c; CES, Center for Epidemiologic Studies

**Table 4b:** Differences on participant characteristics by gender score tertiles.

|  | **Tertile 1**<br>**Masculine**<br>**Characteristics** | **Tertile 2**<br>**Masculine**<br>**and Feminine**<br>**Characteristics** | **Tertile 3**<br>**Feminine**<br>**Characteristics** | **p-value<sup>1</sup>** |
| --- | --- | --- | --- | --- |
| Age in years, mean (SD) | 68.7 (4.8) | 68.2 (5.0) | 68.65 (5.0) | 0.073 |
| Women (N) | 71 | 195 | 277 | <0.001 |
| Men (N) | 277 | 154 | 71 | <0.001 |
| Education, median (IQR) | 16 (6) | 14.5 (6) | 12 (4.5) | <0.001 |
| LDL-C (mg/dl), mean (SD) | 127.8 (35.3) | 131.8 (35.4) | 134.4 (33.7) | 0.044 |
| TC (mg/dl), mean (SD) | 209.5 (40.1) | 216.6 (40.5) | 221.7 (38.1) | <0.001 |
| BMI, mean (SD) | 27.0 (3.8) | 26.9 (4.2) | 26.6 (4.5) | 0.324 |
| Blood pressure, systolic (mmHg), mean (SD) | 144.7 (18.9) | 143.8 (19.5) | 145.8 (71.9) | 0.847 |
| Blood pressure, diastolic (mmHg), mean (SD) | 83.6 (13.1) | 82.9 (11.0) | 87.5 (74.8) | 0.371 |
| Current smoking (N) | 34 | 32 | 30 | 0.876 |
| HbA1c (%), mean (SD) | 5.64 (0.6) | 5.6 (0.6) | 5.5 (0.5) | 0.027 |
| Family history of CVD (N) | 151 (261) | 154 (264) | 166 (272) | n.s. |
| CES-Depression (sum score), median (IQR) | 4 (5) | 6 (7) | 7 (10) | <0.001 |
| Cortisol (nmol/L), mean (SD) | 413.8 (109.1) | 386.7 (114.3) | 370.3 (112.1) | <0.001 |
| Hand grip strength (kg), mean (SD) | 39.7 (9.2) | 33.5 (9.6) | 29.7 (8.4) | <0.001 |
| Negative affect, mean (SD) | 46.6 (8.9) | 48.5 (8.3) | 49.8 (9.8) | <0.001 |
| Life satisfaction, median (IQR) | 53.3 (11.7) | 53.3 (11.7) | 45.5 (15.6) | <0.001 |

<sup>1</sup>Differences over tertiles were assessed using the parametric ANOVA, the nonparametric Kruskal-Wallis test or the $\chi^2$ test where appropriate. Abbreviations: LDL-C, low density lipoprotein-cholesterol; TC, total cholesterol; BMI, body mass index; HbA1c, glycated hemoglobin A1c; CES, Center for Epidemiologic Studies

In a next step, we performed linear regressions to further investigate associations between sex, the GS, and biological and well-being variables. Different linear models were performed to determine the impact of sex and gender separately (Table 5 a) and in combination (Table 5 b). These models revealed that sex was significantly associated with LDL-C and TC, with the GS not additionally showing any association in the models. In contrast, the GS was significantly associated with cortisol levels, CES-depression, negative affect and life satisfaction, with sex not associated with these parameters. Interestingly, hand grip strength was associated with both sex and GS. The associations with hand grip strength, cortisol and life satisfaction were negative suggesting that a more feminine score is associated with lower levels of these parameters, whereas all other parameters were positively associated. That is, independent of sex, a GS associated with more traditionally feminine characteristics was associated with exhibiting lower hand grip strength and cortisol as well as reporting lower levels of life satisfaction. At the same time, independent of sex, a GS associated with more traditionally feminine characteristics was also associated with reporting more depressive symptoms and higher levels of negative affect. HbA1c was not associated with either sex or GS (Table 5b).

**Table 5a:** Association of all significant variables from table 4 /5 by sex and gender score in **separate** linear regression models.

| Dependent variable | Female sex |  |  | Gender Score |  |  |
| --- | --- | --- | --- | --- | --- | --- |
| | $\beta$ | p-value | R<sup>2</sup> | $\beta$ | p-value | R<sup>2</sup> |
| LDL-C | 12.568 | **<0.001** | 0.032 | 0.095 | **0.025** | 0.004 |
| TC | 23.462 | **<0.001** | 0.086 | 0.192 | **<0.001** | 0.014 |
| HbA1c | -0.092 | **0.012** | 0.005 | -0.002 | **0.028** | 0.004 |
| CES-Depression | 1.768 | **<0.001** | 0.022 | 0.060 | **<0.001** | 0.068 |
| Cortisol | -29.088 | **<0.001** | 0.017 | -0.681 | **<0.001** | 0.024 |
| Hand grip strength | -16.100 | **<0.001** | 0.652 | -0.174 | **<0.001** | 0.199 |
| Negative affect | 1.066 | 0.063 | 0.003 | 0.054 | **<0.001** | 0.022 |
| Life satisfaction | -1.938 | **0.001** | 0.01 | -0.108 | **<0.001** | 0.079 |

**Table 5B:**
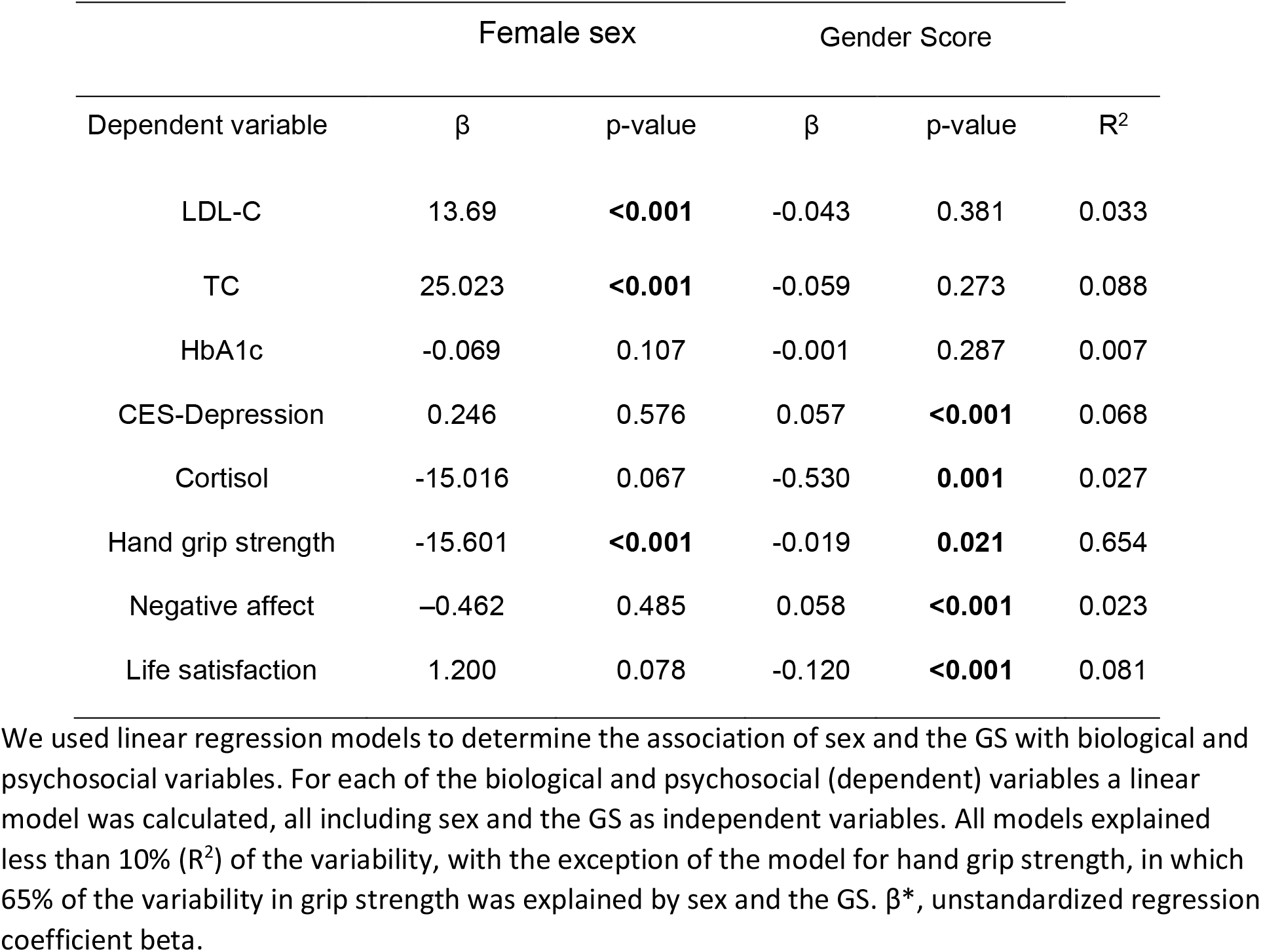
Prediction of all significant variables from table 4 /5 by sex and gender score in **combined** linear regression models.

## Discussion

Our study is the first to construct a GS in a retrospective manner from available study variables. The developed GS was differently distributed in women and men. Biological sex and GS had different predictive power for a number of biological and sociocultural variables, with sex being a better predictor for biological phenotypes and gender being a better predictor for sociocultural ones; for the most part independently of the biological sex of the participants.

We used the BASE-II cohort that provided data on older predominantly healthy adults in and around Berlin, Germany^5^, to construct a GS in analogy to the score that was developed in a mainly Canadian cohort of younger patients with acute coronary syndrome. The novelty of our approach was the fact that we were developing the GS in a completely retrospective manner. BASE-II was primarily not designed to analyze the impact of sex and gender on study data. So, in the primary data assessment in BASE-II, we identified 13 psychosocial and socioeconomic variables that we believed to be associated with the four dimensions of gender^7^. We used these to construct a GS, ranging from 0 (purely masculine) to 100 (purely feminine). The GS of the individual cases in the whole group were almost evenly distributed over the total range from 0 to 100 which was different from the distribution of GS in the only other cohort published so far^1^, where the whole first tertile of patients was clustering in the left corner (GS from 0-10). However, we obtained a similar but not identical frequency distribution of women and men as the previous study, with 55 % of men in the first tertile with more masculine GS and 51 % of women in the 3^rd^ tertile with the more feminine scores^1^. In our cohort, the distributions showed a considerably stronger and more symmetrical overlap with almost a third of women and men in the middle tertile and 13 % of women and 14 % of men having score values of the opposite gender. Thus, the GS gives information clearly different from biological sex, but its distribution is different in the Canadian young diseased and the German healthy aged cohorts.

When constructing the GS, fewer socioeconomic variables than desired were available as may be the case in many clinical and medically focused epidemiological studies. The limited availability of variables may explain differences in the variables that constitute the GS in women and men between our study and previously published data^1^. Furthermore, in the aged cohort of BASE-II, with mainly retired persons, other variables may be gender relevant than in younger cohorts with CVD disease. Psychological parameters, such as perceived stress, loneliness, agreeableness, neuroticism, contributed significantly to the psychosocial differences among women and men.

Furthermore, family status and education, reflecting institutionalized gender, also had a high impact in GS calculation. Family status probably scored high since a large percentage of women in this generation were living as widows or as singles. The influence of education is also well understandable: The traditional role of a daughter in Germany in the 1950’s or earlier, when many of our study participants were born, was to marry, not to go to the university or achieve any higher education.

To assess the impact of GS, we tested the associations of GS with biological variables and with two well-being indicators. Sex was more strongly associated with the more biological variables, i.e. lipid levels and grip strength. Interestingly enough, lipid levels were higher in women, in individuals with female sex and female gender characteristics. This may have been due to chronic undertreatment of high lipid levels in women as reported previously^15, 16^. HbA1c levels were not associated with sex or gender and we assume that the effect of individual treatment may have contributed to this phenomenon.

Gender, in particularly female gender, was more strongly associated with the life style factors, depression, mood-related hormone cortisol, negative effects, and low life satisfaction, than sex. This is in agreement with previously published results^17^.

## Limitations of the study

Our approach has strengths and weaknesses. Limitations result from the retrospective nature of our analysis and the limited number of gender-related variables available in our study. This is, however, an inherent component of our approach: we wanted to develop a method that enables researchers to measure gender in a retrospective manner since we feel that many studies exist in which an estimate of gender, even if retrospective, could uncover new aspects and reveal important insights.

Second, we chose to use reported sex as a dependent variable in our logistic regression analysis because there is no gold standard for the gender, reported sex is likely the closest measure of gender available in most surveys. BEM sex role inventory could have been used as a dependent variable instead of the sex. However, this scale only includes personality traits that are only encompass the gender identity aspect of the gender construct and was not present in our retrospective dataset.

## Conclusions and future aspects

With our strategy, we were able to develop a gender score (GS) in a retrospective manner from available study variables that characterized women and men in addition to biological sex. This means that the notion of gender can be retrospectively introduced into a large number of studies. In future approaches, we will test how sex and the gender score respectively predict clinical outcomes in this predominantly healthy cohort. If we find that the gender score acts as an outcome predictor beyond clinical parameters, new preventive options may arise.

## Data Availability

Data can be made available under the legal guidelines of the stated institute when needed.

## Abbreviations

ACS: Acute Coronary Syndrome
BASE-II: Berliner Aging Study II
BFI: Big Five Inventory personality traits scale
CES-D: Center for Epidemiologic Studies Depression Scale
CVD: Cardiovascular diseases
GS: Gender Score
HbA1c: Hemoglobin A1c,
LDL: low-density lipoproteins
LRA: Logistic Regression Analysis
PCA: Principal Component Analysis
PSS: Perceived Stress Scale
TC: Total Cholesterol
TICS: Trier Inventory for Chronic Stress
UCLA-Loneliness: University of California, Los Angeles Loneliness Scale

## Acknowledgements

### Funding

The current work was part of the study GendAge (https://gendage.charite.de/en/) supported by grants from the German Federal Ministry of Education and Research (Bundesministerium für Bildung und Forschung, BMBF) to Ilja Demuth and Vera Regitz-Zagrosek (grant number #01GL1716A) and Denis Gerstorf (#01GL1716B). Dr Pilote holds a James McGill chair at McGill University and was the recipient of the Berlin Institute of Health’s Excellence Award for Sex and Gender Aspects in Health Research that supported in part this work. This article is also based on data from the Berlin Aging Study II (BASE-II, Co-PIs are Lars Bertram, Ilja Demuth, Denis Gerstorf, Ulman Lindenberger, Graham Pawelec, Elisabeth Steinhagen-Thiessen, and Gert G. Wagner). BASE-II was supported by the German Federal Ministry of Education and Research (Bundesministerium für Bildung und Forschung, BMBF) under grant numbers #01UW0808; #16SV5536K, #16SV5537, #16SV5538 and #16SV5837. Another significant source of funding was the Max Planck Institute for Human Development, Berlin, Germany.

## Notes

### Competing Interest Statement

The authors have declared no competing interest.

